# Theoretical framework for retrospective studies of the effectiveness of SARS-CoV-2 vaccines

**DOI:** 10.1101/2021.01.21.21250258

**Authors:** Joseph A. Lewnard, Manish M. Patel, Nicholas P. Jewell, Jennifer R. Verani, Miwako Kobayashi, Mark Tenforde, Natalie E. Dean, Benjamin J. Cowling, Benjamin A. Lopman

## Abstract

Observational studies of the effectiveness of vaccines to prevent COVID-19 are needed to inform real-world use. These are now in planning amid the ongoing rollout of SARS-CoV-2 vaccines globally. While traditional case-control (TCC) and test-negative design (TND) studies feature prominently among strategies used to assess vaccine effectiveness, such studies may encounter important threats to validity. Here we review the theoretical basis for estimation of vaccine direct effects under TCC and TND frameworks, addressing specific natural history parameters of SARS-CoV-2 infection and COVID-19 relevant to these designs. Bias may be introduced by misclassification of cases and controls, particularly when clinical case criteria include common, non-specific indicators of COVID-19. When using diagnostic assays with high analytical sensitivity for SARS-CoV-2 detection, individuals testing positive may be counted as cases even if their symptoms are due to other causes. The TCC may be particularly prone to confounding due to associations of vaccination with healthcare-seeking behavior or risk of infection. The TND reduces but may not eliminate this confounding, for instance if individuals who receive vaccination seek care or testing for less-severe infection. These circumstances indicate the two study designs cannot be applied naively to datasets gathered through public health surveillance or administrative sources. We suggest practical strategies to reduce bias in vaccine effectiveness estimates at the study design and analysis stages.

## BACKGROUND

Non-randomized studies undertaken after vaccine authorization or licensure provide crucial information about real-world vaccine effectiveness (VE). Such studies may also address questions not answered by clinical trials, such as the duration of protection, the effectiveness of alternative dosing schedules, and the level of protection achieved against emerging variants or within various population subgroups.^1,2^ While prospective cohort studies provide an opportunity to compare outcomes among vaccinated and unvaccinated individuals, such studies may be logistically prohibitive due to the large size and complexity of longitudinal follow-up. Thus, retrospective studies comparing prior vaccination among individuals with a known clinical outcome of disease or no disease (i.e. case-control VE studies) provide an efficient alternative.^3^

Phase III randomized controlled trials (RCTs) have established efficacy of multiple vaccines against COVID-19,^4–6^ with others in progress. Observational VE studies have been prioritized by public health^7,8^ and regulatory^9^ authorities to inform real-world use. Traditional case-control (TCC) and test-negative design (TND) studies are the primary retrospective frameworks for assessing VE,^10^ and plans to use such study designs to assess effectiveness of SARS-CoV-2 vaccines are being developed.^7^ However, these and other observational designs may differ with respect to measures of VE and risks of bias.^2^

Here we lay out the theoretical basis for use of the TCC and TND in assessments of vaccine direct effects against SARS-CoV-2 infection and COVID-19. We highlight key assumptions and potential biases underlying VE estimates from the TCC and TND studies, and provide practical recommendations to guide researchers through stages of study design, analysis, and interpretation.

## OVERVIEW OF THE DESIGNS

The TCC and TND derive VE estimates by comparing the odds of prior vaccination among individuals who, at the time of enrollment, are cases (experiencing the clinical endpoint against which VE is to be measured) or non-cases (controls). Cases are typically individuals who meet clinical criteria such as the presence of predefined symptoms together with laboratory-confirmed detection of a vaccine-targeted infectious agent.^11^ Under the TCC, controls are selected from a pool of individuals who are not known to be experiencing the clinical endpoint of interest at the time of enrollment, but are members of the same population from which cases are identified. Such individuals may be community controls, selected from among asymptomatic individuals in the community; registry controls, selected from population-based registries; or healthcare controls, selected among patients experiencing an alternate disease unrelated to the pathogen and vaccine of interest.^12^

The TND is an alternative to the TCC that has become the default approach for studies of vaccines against influenza, rotavirus, and other pathogens.^13^ In the TND, both cases and controls are selected from among individuals who receive diagnostic tests for a pathogen of interest; conventionally, individuals are tested because they experience a clinical syndrome which may be caused by the vaccine-targeted pathogen or other agents. Cases are those testing positive, and controls are those testing negative, placing a heavy reliance on diagnostic accuracy.^2,14,15^ Since the TND does not require active selection of controls from the community or other settings, it may offers logistical advantages over the TCC.

Below we lay out the general framework for estimation of vaccine direct effects in TCC and TND studies. We will introduce relevant notation and assumptions and then extend this framework to address observations that may be expected in studies of SARS-CoV-2.

## IDENTIFICATION OF VACCINE EFFECTIVENESS UNDER TCC AND TND DESIGNS

Consider the instantaneous risk (or prevalence) of a clinical condition of interest to be *p* (in an unvaccinated population), a value between 0 and 1 (**Table 1**). Consider that vaccination reduces individuals’ risk of the condition by a factor VE= (1 − *θ*), such that the relative risk of the condition for vaccinated (versus unvaccinated) individuals is *θ*; we define 1 − *θ* as the vaccine “direct effect” resulting from reduction in biological susceptibility to the condition of interest due to vaccination. Vaccine protection could be achieved through differing means, such as uniformly reducing individuals’ risk by a factor of (1 − *θ*), or by preventing the condition with certainty among a proportion (1 − *θ*)of recipients while failing to confer any protection among others. Unless otherwise noted in our summary, these differing mechanisms of vaccine action (elsewhere termed “leaky” or “all-or-nothing” protection^10^, respectively) do not result in practical differences in our analyses of VE.

**Table 1:**
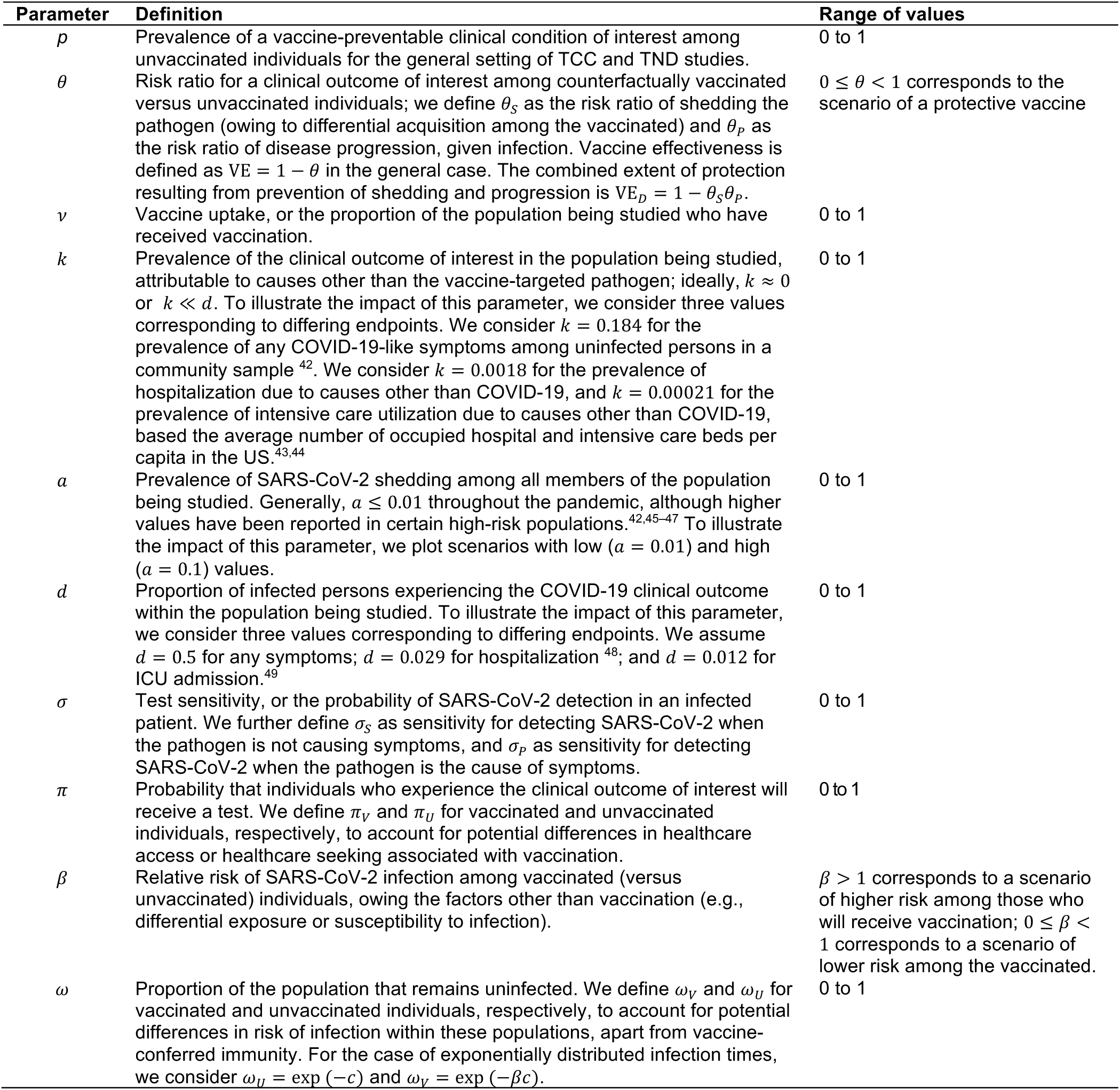
Parameters and their definitions.

| Parameter | Definition | Range of values |
| --- | --- | --- |
| $p$ | Prevalence of a vaccine-preventable clinical condition of interest among unvaccinated individuals for the general setting of TCC and TND studies. | 0 to 1 |
| $\theta$ | Risk ratio for a clinical outcome of interest among counterfactually vaccinated versus unvaccinated individuals; we define $\theta_s$ as the risk ratio of shedding the pathogen (owing to differential acquisition among the vaccinated) and $\theta_p$ as the risk ratio of disease progression, given infection. Vaccine effectiveness is defined as $VE = 1 - \theta$ in the general case. The combined extent of protection resulting from prevention of shedding and progression is $VE_D = 1 - \theta_s \theta_p$. | $0 \leq \theta < 1$ corresponds to the scenario of a protective vaccine |
| $v$ | Vaccine uptake, or the proportion of the population being studied who have received vaccination. | 0 to 1 |
| $k$ | Prevalence of the clinical outcome of interest in the population being studied, attributable to causes other than the vaccine-targeted pathogen; ideally, $k \approx 0$ or $k \ll d$. To illustrate the impact of this parameter, we consider three values corresponding to differing endpoints. We consider $k = 0.184$ for the prevalence of any COVID-19-like symptoms among uninfected persons in a community sample<sup>42</sup>. We consider $k = 0.0018$ for the prevalence of hospitalization due to causes other than COVID-19, and $k = 0.00021$ for the prevalence of intensive care utilization due to causes other than COVID-19, based the average number of occupied hospital and intensive care beds per capita in the US.<sup>43,44</sup> | 0 to 1 |
| $a$ | Prevalence of SARS-CoV-2 shedding among all members of the population being studied. Generally, $a \leq 0.01$ throughout the pandemic, although higher values have been reported in certain high-risk populations.<sup>42,45–47</sup> To illustrate the impact of this parameter, we plot scenarios with low ($a = 0.01$) and high ($a = 0.1$) values. | 0 to 1 |
| $d$ | Proportion of infected persons experiencing the COVID-19 clinical outcome within the population being studied. To illustrate the impact of this parameter, we consider three values corresponding to differing endpoints. We assume $d = 0.5$ for any symptoms; $d = 0.029$ for hospitalization<sup>48</sup>; and $d = 0.012$ for ICU admission.<sup>49</sup> | 0 to 1 |
| $\sigma$ | Test sensitivity, or the probability of SARS-CoV-2 detection in an infected patient. We further define $\sigma_s$ as sensitivity for detecting SARS-CoV-2 when the pathogen is not causing symptoms, and $\sigma_p$ as sensitivity for detecting SARS-CoV-2 when the pathogen is the cause of symptoms. | 0 to 1 |
| $\pi$ | Probability that individuals who experience the clinical outcome of interest will receive a test. We define $\pi_v$ and $\pi_u$ for vaccinated and unvaccinated individuals, respectively, to account for potential differences in healthcare access or healthcare seeking associated with vaccination. | 0 to 1 |
| $\beta$ | Relative risk of SARS-CoV-2 infection among vaccinated (versus unvaccinated) individuals, owing the factors other than vaccination (e.g., differential exposure or susceptibility to infection). | $\beta > 1$ corresponds to a scenario of higher risk among those who will receive vaccination; $0 \leq \beta < 1$ corresponds to a scenario of lower risk among the vaccinated. |
| $\omega$ | Proportion of the population that remains uninfected. We define $\omega_v$ and $\omega_u$ for vaccinated and unvaccinated individuals, respectively, to account for potential differences in risk of infection within these populations, apart from vaccine-conferred immunity. For the case of exponentially distributed infection times, we consider $\omega_u = \exp(-c)$ and $\omega_v = \exp(-\beta c)$. | 0 to 1 |

Defining *v* as the proportion of the population receiving vaccine, the probabilities of individuals in the population falling into various case and control categories, given their vaccination status, is laid out in the two-by-two tables in **Table 2**. Under the TCC,

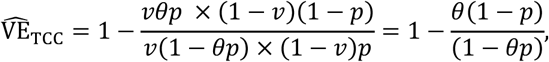

which, for rare outcomes (*p* ≈ 0), yields an unbiased estimate 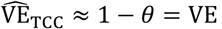 Alternative design approaches such as incidence density matching of cases and controls,^16^ inclusive sampling,^17^ or other strategies may reduce reliance on the rare outcome assumption; here for simplicity we focus on more general TCCs designs.

**Table 2:** Two-by-two tables for TCC and TND studies.

| Design | Exposure | Outcome |  |
| --- | --- | --- | --- |
| Traditional case control |  | <u>Case</u> | <u>Control</u> |
| | Vaccinated | $v\theta p$ | $v(1 - \theta p)$ |
| | Unvaccinated | $(1 - v)p$ | $(1 - v)(1 - p)$ |
| Test negative design |  | <u>Test-positive</u> | <u>Test-negative</u> |
| | Vaccinated | $v\theta p$ | $vk$ |
| | Unvaccinated | $(1 - v)p$ | $(1 - v)k$ |

Under the TND, controls instead experience an outcome that is unaffected by vaccination against the pathogen of interest.^14,15,18^ Defining the risk of this outcome as *k*, regardless of vaccination (**Table 2**),

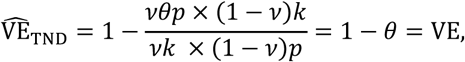

which does not require the rare-disease assumption to recover the vaccine direct effect.

## CLASSES OF VACCINE DIRECT EFFECTS

Vaccine-conferred protection may impact multiple aspects of the natural history of infectious disease agents such as SARS-CoV-2, including but not limited to prevention of virus acquisition, reduced viral replication in the upper respiratory mucosa, earlier clearance of infection, and prevention of mild or severe symptoms. The differential effects of SARS-CoV-2 vaccines against infection and disease endpoints remain imperfectly understood^19^ and may vary by vaccine product and class.

The typical estimands of interest in TCC and TND studies are VE against symptomatic disease of any severity, or VE against severe disease. These are also typical target estimands in individually-randomized vaccine efficacy trials.^20^ Consider *θ*_*s*_ to be the relative risk of infection for a vaccinated versus unvaccinated individual, which may result from reductions in host susceptibility to acquisition of SARS-CoV-2 or accelerated clearance of the pathogen.^20^ Consider *θ*_*p*_ to be the relative risk of COVID-19 (or COVID-19 meeting a particular severity threshold, e.g. hospitalization, intensive care unit admission, or mechanical ventilation) for an infected vaccinated individual versus an infected unvaccinated individual. Protection resulting from prevention of both infection and progression is then VE_*D*_=1− *θ*_*s*_*θ*_*p*_; we assume VE_*D*_ is the desired estimand and describe biases of 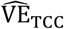 and 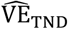 relative to VE_*D*_. Specialized designs would be required to isolate *θ*_*s*_ or *θ*_*p*_.

## OUTCOME MISCLASSIFICATION

### Etiologic detection

Clinical outcomes of interest to researchers may range from laboratory confirmation of infection regardless of symptoms^21–23^ to laboratory-confirmed mild, moderate, or severe disease manifestations.^20^A positive test result may not specifically indicate the etiology of current symptoms, as SARS-CoV-2 genetic material can be shed for prolonged periods after infection.^24^ Moreover, many acquisitions of SARS-CoV-2 may never lead to symptoms.^25^

Prolonged viral shedding and the presence of asymptomatic infection may introduce misclassification in both TCC and TND studies. For instance, TCC studies enrolling controls selected from the community, from registries, or among patients seeking care for other (e.g., non-respiratory) diseases, may suffer from misclassification if a proportion of these controls are in fact infected with SARS-CoV-2, but not tested due to the absence of symptoms. Inclusion of infected individuals in the control group will diminish the apparent effects of vaccination, provided vaccine prevents infection. Likewise, in both TCC and TND studies, individuals who are shedding SARS-CoV-2 genetic material but experiencing illness due to another cause (e.g., another infection or a non-infectious process with overlapping symptoms such as chronic obstructive pulmonary disease) may be misclassified as cases. Misclassification of asymptomatically infected persons as controls, and of infected symptomatic persons as cases when their illness is not caused by SARS-CoV-2, can be expected to increase with higher prevalence of infection in the community.

To formalize resulting biases, consider separately the risks of SARS-CoV-2 acquisition and disease progression. Defining *a* as the instantaneous risk (prevalence) of being infected with SARS-CoV-2, at any time, and *d* as the instantaneous risk of experiencing symptoms, given infection, the risk of symptomatic and asymptomatic SARS-CoV-2 infections are *ad* and *a*(1 − *d*), respectively. We extend our contingency tables to accommodate test-positive symptomatic cases, asymptomatic controls, and test-negative symptomatic controls in **Table 3**. Here, the TCC yields

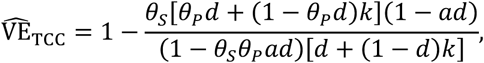

which remains subject to bias owing to the risk of symptoms due to causes other than SARS-CoV-2 (*k*) and the prevalence of SARS-CoV-2 infection (*a*). However, these biases may be ignorable under several conditions. If prevalence of other conditions is low or negligible (*k* ≈ 0, as may be true for very severe endpoints), or negligible relative to the risk of COVID-19 for an infected individual (*k* ≪ *d*, if endpoints are chosen with high specificity for COVID-19), the TCC yields

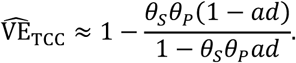

**Table 3:**
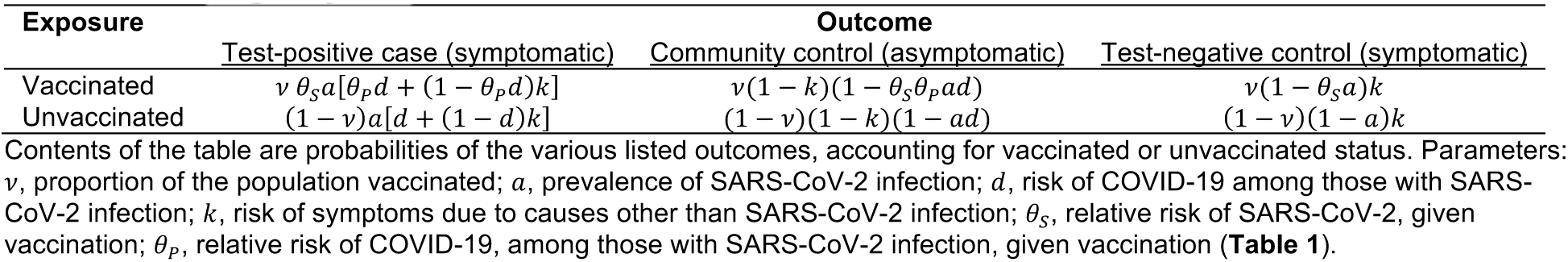
Contingency tables for TCC and TND studies with outcome misclassification.

| Exposure | Outcome |  |  |
| --- | --- | --- | --- |
|  | Test-positive case (symptomatic) | Community control (asymptomatic) | Test-negative control (symptomatic) |
| Vaccinated | $v \theta_s a [\theta_p d + (1 - \theta_p d)k]$ | $v(1 - k)(1 - \theta_s \theta_p a d)$ | $v(1 - \theta_s a)k$ |
| Unvaccinated | $(1 - v)a[d + (1 - d)k]$ | $(1 - v)(1 - k)(1 - a d)$ | $(1 - v)(1 - a)k$ |

Such a design may under-estimate VE_*D*_ if prevalence of SARS-CoV-2 infection (*a*) is high owing to inclusion of infected asymptomatic individuals in the control group. If prevalence of SARS-CoV-2 infection in the community is low or negligible (*a* ≈ 0). 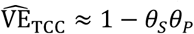

Comparing symptomatic individuals who test positive and negative for infection, the TND yields

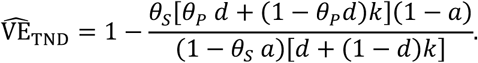

The same conditions by which bias can be reduced under the case-control design reduce bias under the test-negative design. With *k* ≈ 0 or *k* ≪ *d*,

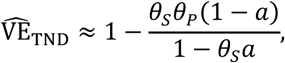

and with (*a* ≈ 0), 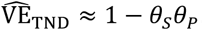

We illustrate the quantitative extent of bias under the TCC and TND in **Figure 1**. Under most realistic situations, including a highly efficacious vaccine (VE_*D*_∼95%), low prevalence of acute infection (*a* < 0.05), and substantially increased risk of symptoms, given infection (*d* ≫ *k*), bias is low or negligible for both designs. While relatively insensitive to changes in prevalence of infection within ranges that have been reported for SARS-CoV-2, bias increases if the risk of disease, given infection, is low (i.e. if non-specific or prevalent symptoms are included in clinical case criteria), potentially leading to substantial under-estimation of VE_*D*_.

**Figure 1:**
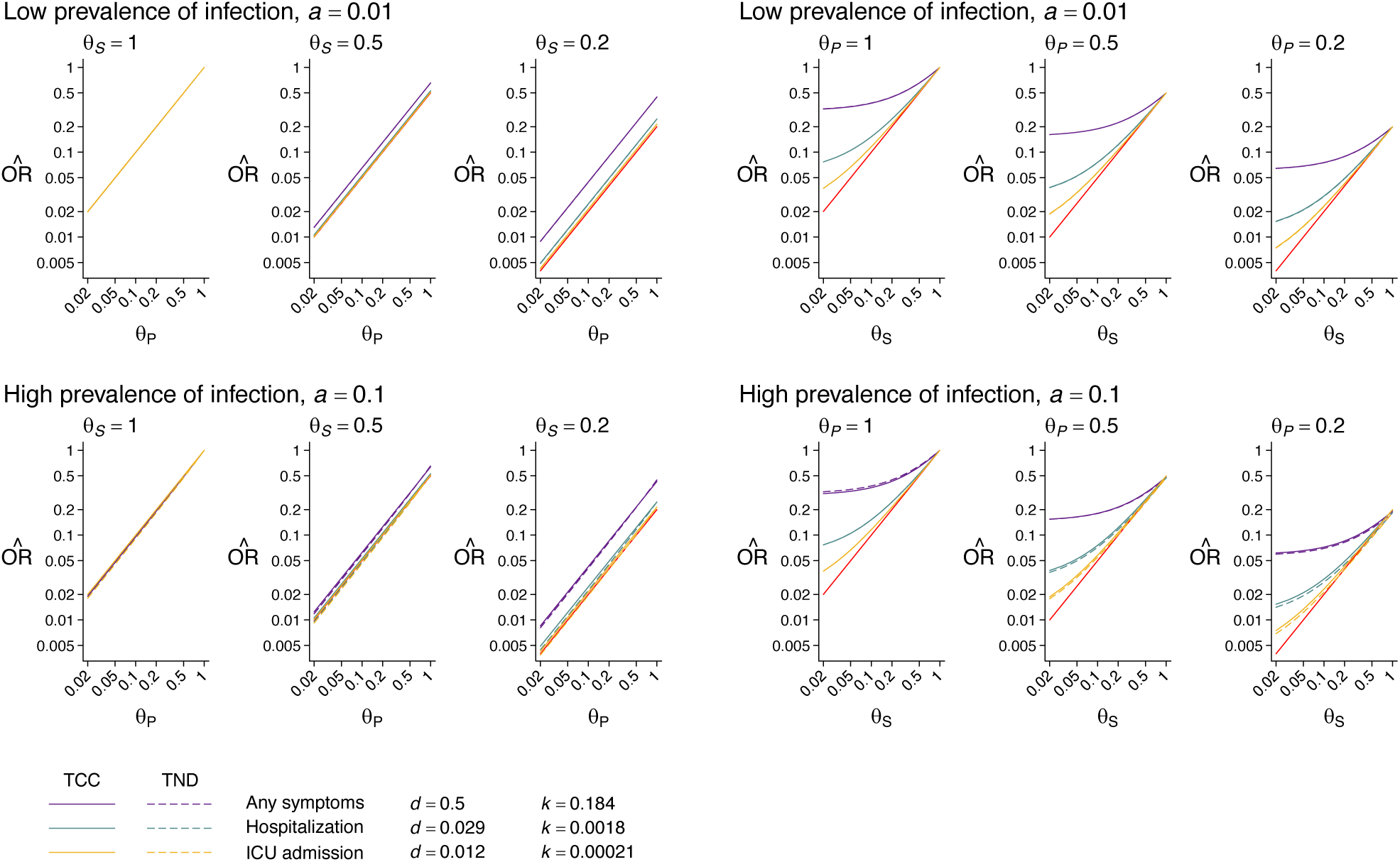
True and estimated vaccine effectiveness under the TCC and TND. We illustrate expected estimates of the odds ratio of vaccination given case versus control status under the TCC (solid lines) and TND (dashed lines) with differing prevalence of infection (top row, *a =* 0.01; bottom row, *a =* 0.1). Panels on the left and right illustrate effect measures with varying values of *θ* _*p*_ (for fixed values of *θ*_*s*_) and *θ*_*s*_ (for fixed values of *θ*_*p*_), respectively. Colors correspond to values of *d* and *k* for differing endpoints based on the estimates laid out in **Table 1**. Red diagonal lines across each panel illustrate the true effect.

### Choice of diagnostic assays

Multiple assays are available for SARS-CoV-2 detection, and the use of assays with differing performance characteristics may influence the extent of biases identified above. Nucleic acid amplification assays such as polymerase chain reaction (PCR), for instance, have higher analytic sensitivity (i.e., likelihood of positive test for sample containing viral material at the minimum detectable concentration for which assay is designed), potentially leading to detection of SARS-CoV-2 genetic material among individuals with low levels of virus shedding. Antigen detection tests, with lower analytic sensitivity, may in contrast yield negative results for individuals with low virus shedding that is detectable by molecular testing.^26^ In comparison to mild illness, both assays may have lower sensitivity for severe disease which can be delayed in presentation, occurring often during the second week after infection onset when viral shedding is lower.^42^

Define *σ*_*p*_ and *σ*_*s*_ as the sensitivity of an assay for detecting SARS-CoV-2 among individuals experiencing COVID-19 and those not experiencing symptoms attributable to SARS-CoV-2, respectively. We limit our consideration to differences in test sensitivity, considering specificity to encompass the related issue of non-etiologic SARS-CoV-2 detections; under field conditions, antigen and molecular tests have been found to have similar analytical specificity.^26,27^ The limiting case of an assay which perfectly rules out SARS-CoV-2 detections that are not causing symptoms (*σ*_*s*_ = 0) yields the contingency table presented in **Table 4** for case and control groups (**Table S1** extends this for non-zero values of *σ*_*s*_).

**Table 4:** Contingency tables for TCC and TND studies with exclusion of SARS-CoV-2 shedding from COVID-19.

| Exposure | Outcome |  |  |
| --- | --- | --- | --- |
|  | Test-positive case (symptomatic) | Community control (asymptomatic) | Test-negative control (symptomatic) |
| Vaccinated | $v \theta_s \theta_p a d \sigma_p$ | $v(1-k)(1-\theta_s \theta_p a d)$ | $v[k + (1-k)\theta_s \theta_p a d(1-\sigma_p)]$ |
| Unvaccinated | $(1-v)a d \sigma_p$ | $(1-v)(1-k)(1-a d)$ | $(1-v)[k + (1-k)a d(1-\sigma_p)]$ |

With *σ*_*s*_ = 0, the TCC yields

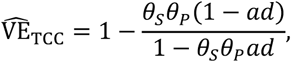

removing the reliance on assumptions of *k* ≈ 0 or *k* ≪ *d*, and allowing 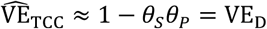 if overall prevalence of infection is low (*a* ≈ 0). Under the TND,

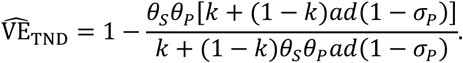

Here, we obtain an unbiased estimate 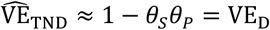 only with high sensitivity of the assay for individuals experiencing COVID-19 (*σ*_*p*_ ≈ 1). Lower values of *σ*_*p*_ result in greater bias (**Figure 2**). Nonetheless, under a scenario where *σ*_*s*_ is high and asymptomatic SARS-CoV-2 detections are minimal (*σ*_*p*_ ≈ 0; **Figure 2**), the extent of bias remains lower than what arises with perfect detection of SARS-CoV-2 in both asymptomatic infection and COVID-19 (as plotted in **Figure 1**), particularly for vaccines that primarily prevent infection (**Figure 2**).

**Figure 2:**
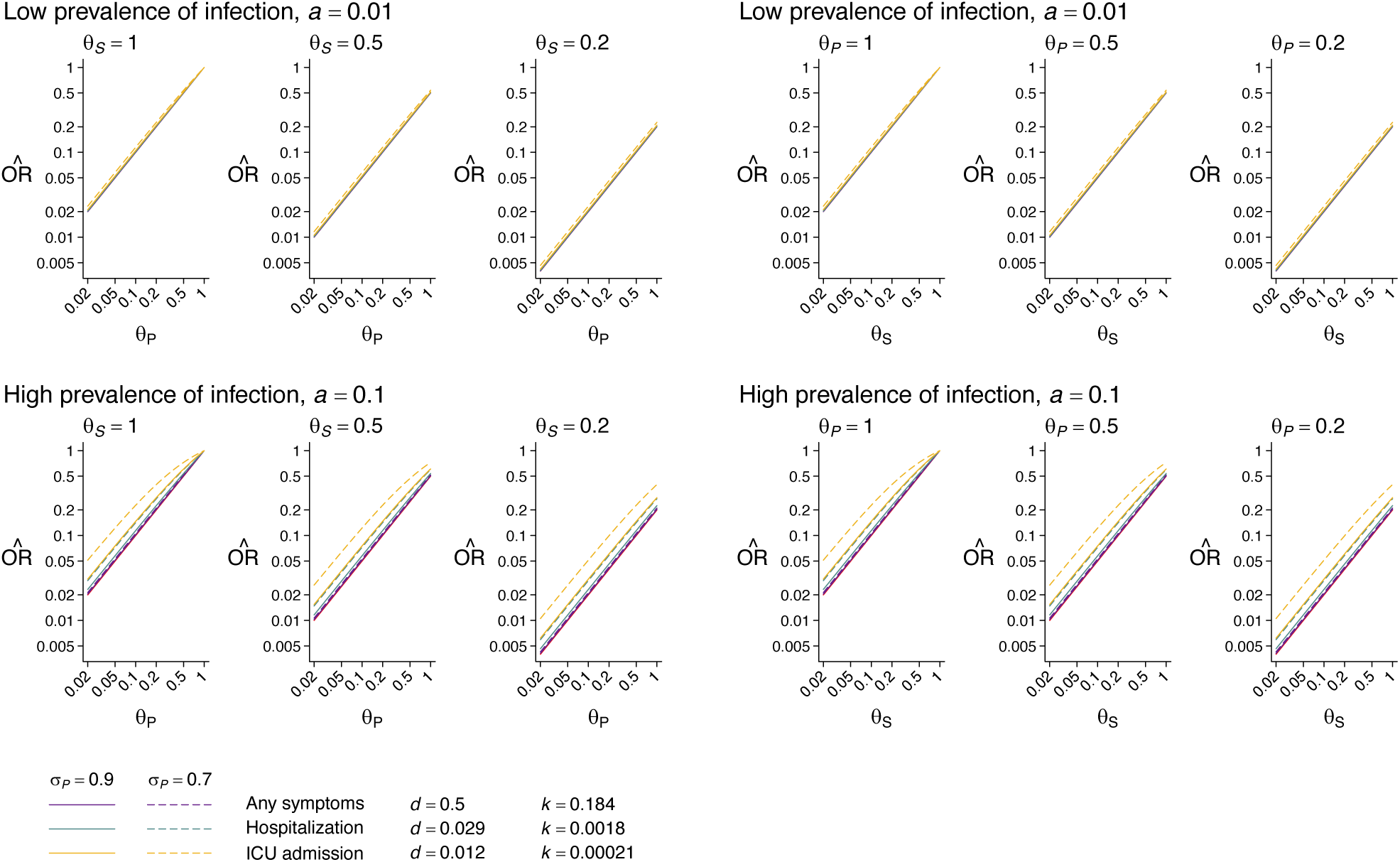
Estimated vaccine effectiveness under the TND with discrimination of non-etiologic SARS-CoV-2 detection. We illustrate expected estimates of the odds ratio of vaccination given case versus control (test-negative) status assuming *σ*_*s*_ *=* 0, under conditions with differing sensitivity for SARS-CoV-2 detection among COVID-19 cases (*σ*_*p*_ *=*0.9, solid lines; *σ*_*p*_*=*0.7, dashed lines), considering differing prevalence of infection (top row, *a =* 0.01; bottom row, *a =* 0.1). Panels on the left and right illustrate effect measures with varying values of *θ*_*p*_ (for fixed values of *θ*_*s*_) and *θ*_*s*_ (for fixed values of *θ*_*p*_), respectively. Colors correspond to values of *d* and *k* for differing endpoints based on the estimates laid out in **Table 1**. Red diagonal lines across each panel illustrate the true effect.

We note that no available assay enables perfect differentiation of COVID-19 from non-etiologic detections of SARS-CoV-2; statistical corrections for imperfect test sensitivity and specificity have been proposed to adjust final VE estimates.^28^ Further complicating the selection of assays, available data suggest similar viral loads and/or nucleic acid abundance among individuals with or without symptoms at similar stages of infection.^29,30^ However, onset of COVID-19 upper respiratory tract symptoms typically coincides with peak viral shedding. Thus, individuals seeking care for new-onset COVID-19 may generally have higher viral load than individuals experiencing symptoms owing to other causes, who may be at later stages of SARS-CoV-2 shedding or molecular detection.^31^ This circumstance may drive an artefactual association of higher SARS-CoV-2 shedding with COVID-19.

## HEALTHCARE-SEEKING BEHAVIOR

Differential healthcare-seeking behaviors of individuals who can access and receive vaccines, versus others, may be another source of bias. The TND aims to limit such bias by restricting enrollment to individuals who seek care and receive diagnostic testing for the same or similar clinical indications. In TCC studies, there may be uncertainty about whether controls were truly disease-free, as some may not have sought care given symptoms. This may be particularly true if studies select controls from registries without direct outreach.

We may quantify the resulting bias by considering the number of controls who are misclassified when the probability of seeking care, given symptoms, is *π*_*v*_ among the vaccinated and *π*_*u*_ among the unvaccinated. Defining eligible controls as individuals who do not become ascertained as cases (**Table 5**):

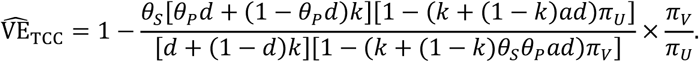

**Table 5:**
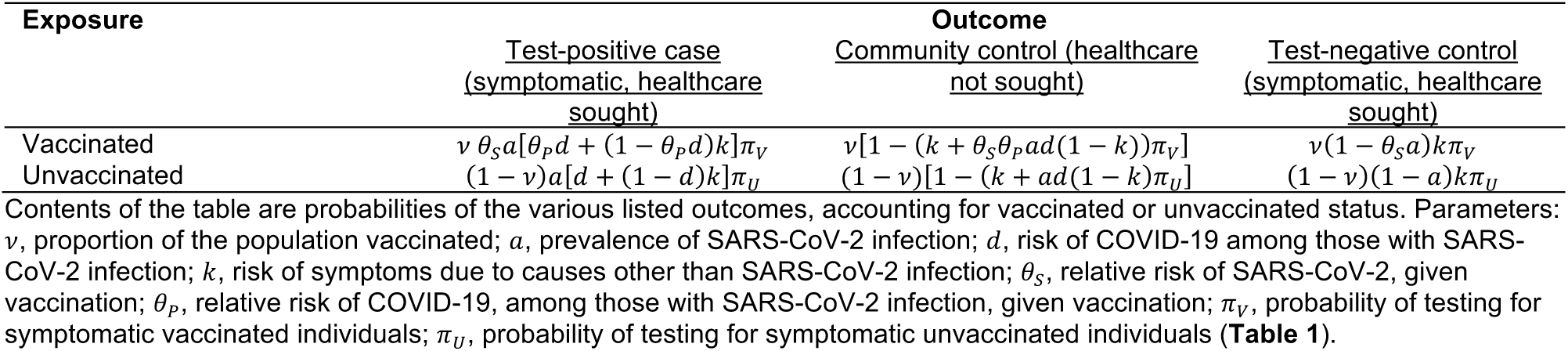
Contingency table for TCC and TND studies with differential healthcare-seeking among the vaccinated and unvaccinated.

The resulting bias generally leads to under-estimation of true protection (VE_D_)when the probability of seeking treatment is higher among vaccinated than unvaccinated persons, given the same status of being infected or symptomatic, owing to the disproportionately higher likelihood for vaccinated individuals to be ascertained as cases (**Figure 3**). However, studies enrolling health facility controls may encounter further difficulty. If vaccination is associated with higher rates of healthcare seeking, prevalence of vaccination among controls seeking care for less-severe conditions may be inflated, leading to overestimation of VE_D_.

**Figure 3:**
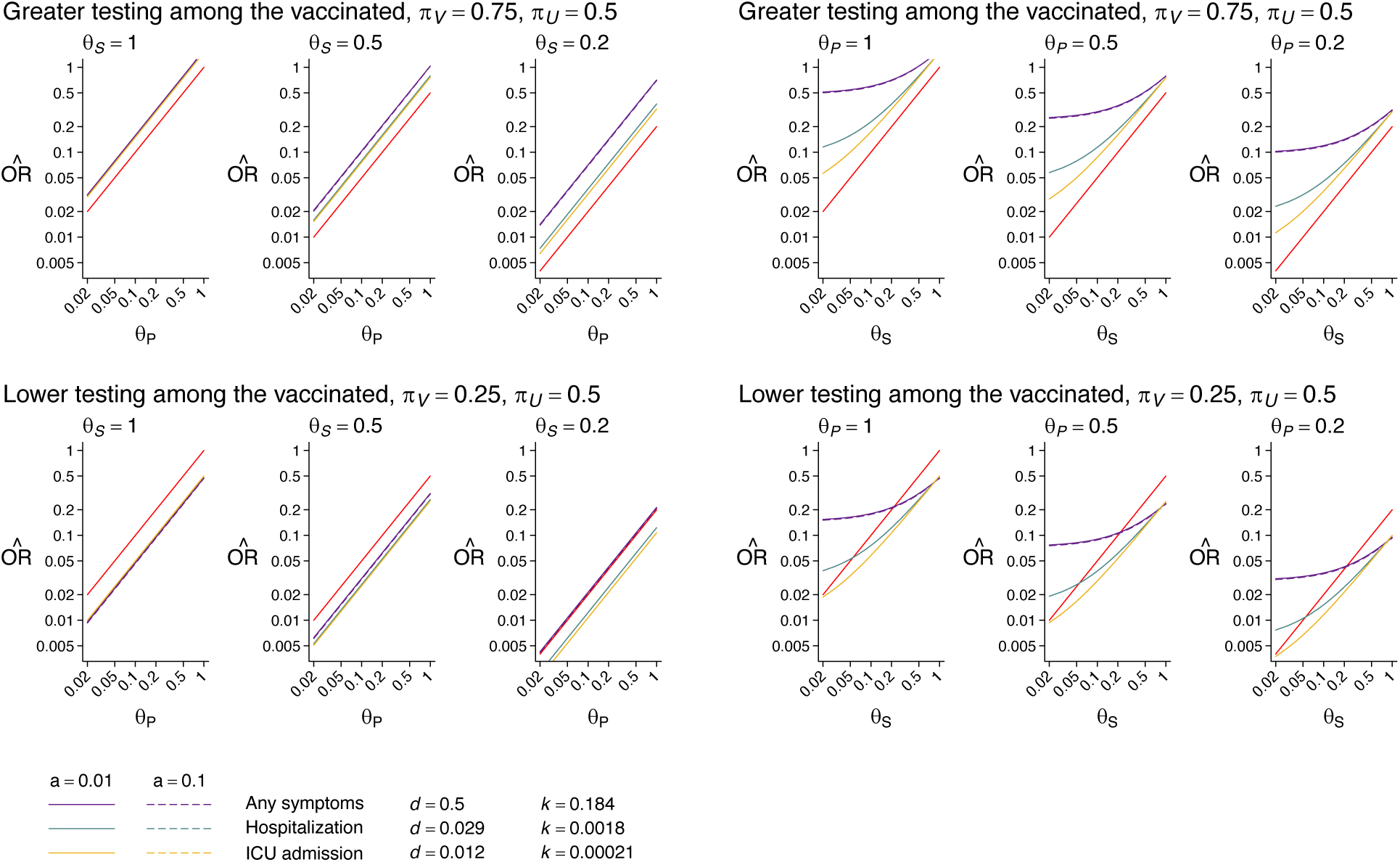
Estimated vaccine effectiveness under the TCC with differential testing, given symptoms, among vaccinated and unvaccinated persons. We illustrate expected estimates of the odds ratio of vaccination given case versus control status, considering differing likelihood of testing among vaccinated and unvaccinated individuals, given the same clinical presentation (top row, greater testing among the vaccinated; bottom row, lower testing among the vaccinated). Panels on the left and right illustrate effect measures with varying values of *θ*_*p*_ (for fixed values of *θ*_*s*_) and *θ*_*s*_ (for fixed values of *θ*_*p*_), respectively. Colors correspond to values of *d* and *k* for differing endpoints based on the estimates laid out in **Table 1**; solid and dashed lines correspond to low (*a =* 0.01) and high (*a =* 0.1) infection prevalence scenarios, respectively. Red diagonal lines across each panel illustrate the true effect.

Applying the same formulation under the TND, the terms *π*_*v*_ and *π*_*u*_ cancel out (**Table 5**). Nonetheless, in practical contexts, TND studies may enroll individuals across a spectrum of clinical severity. This may cause bias to persist if the probability of treatment-seeking differs for vaccinated and unvaccinated individuals with relatively less severe or more severe illness.^18^ These considerations motivate approaches such as matching of cases and test-negative controls (or cases and alternative-disease controls under the TCC) on clinical severity.

Prevalence of other causes of illness leading to care-seeking (e.g., among alternative-disease controls under the TCC and among test-negative controls under the TND) may vary considerably over time. If risk of disease due to these other causes is also associated with likelihood of vaccination, further bias may arise. Even under normal circumstances, disease etiology among controls can change within a season or from year to year. This variation is likely to be extreme over the time horizon that SARS-CoV-2 vaccines are rolled out, as non-pharmaceutical interventions (NPIs) targeting SARS-CoV-2 have affected circulation of other pathogens.^32,33^ As NPIs are rolled back, other causes of healthcare-seeking may surge. Since many infections causing respiratory symptoms (e.g., influenza, pneumococcus) are vaccine preventable, controls with these infections may be less likely to have sought vaccine. Accordingly, the ratio *π*_*v*_ /*π*_*u*_ may change over time.

## A CAUTIONARY NOTE

Testing for SARS-CoV-2 is commonly undertaken for both clinical diagnostic and public health purposes. Thus, data on SARS-CoV-2 test results may encompass individuals tested because of clinical symptoms as well as those tested due to contact with a known or suspected COVID-19 case, for purposes of screening before work or medical procedures, or for other indications. Stratifying analyses to address fairly homogeneous populations, or to individuals who receive SARS-CoV-2 testing with the same frequency or for the same indications, may be of considerable importance.

Variation in the likelihood of seeking testing is expected to be greatest for cases of mild illness or in the absence of any symptoms. There may be substantial association with individuals’ healthcare utilization, including the propensity to be vaccinated. In the most extreme case, populations targeted for vaccination may also be populations targeted for frequent testing, such as healthcare providers, residents of long-term care facilities, and essential workers. Infections in these groups will be more likely to be detected relative to infections within groups without similar access to testing, who may also be less likely to be vaccinated. Here *π*_*v*_ ≫ *π*_*u*_, potentially leading to substantial under-estimation of VE. Similar bias may persist when vaccines are made more widely available if individuals with the greatest propensity to seek healthcare, given the same clinical presentation, including SARS-CoV-2 testing, are also more likely to be vaccinated.

Thus, the TND cannot be applied naively to comprehensive SARS-CoV-2 testing datasets gathered through public health surveillance or administrative data sources. To adequately correct for potential differences in vaccinated and unvaccinated individuals’ likelihood of being tested, designs or analyses should compare individuals being tested for the same indications (e.g., fulfilling particular clinical criteria, known high-risk exposure to SARS-CoV-2, or routine screening), for instance by matching or stratifying on occupation. Bias may be further reduced in studies considering severe disease endpoints for which both vaccinated and unvaccinated individuals would be unlikely to defer treatment, regardless of their propensity to seek healthcare for less severe illness.

## ASSOCIATION OF VACCINE UPTAKE WITH DIFFERENTIAL RISK OF INFECTION

### Confounding in prevalence of infection and immunity

Limited vaccine supplies are being targeted at groups most likely to be exposed to SARS-CoV-2 as well as those at greatest risk of severe COVID-19, if infected. Even when vaccines are more widely available, uptake of vaccination may be highest among individuals who adhered most strongly to NPIs. Accordingly, exposure to infection and risk of progressing to severe disease will not be equal between vaccinated and unvaccinated groups. This differential extends not only to rates of new infection, but also to the prevalence of prior natural infection and immunity. As available evidence suggests naturally acquired immunity is strongly (∼90%) protective against re-infection, there may be substantial differences in risk among vaccinated and unvaccinated individuals owing to factors other than vaccine-derived protection.^34,35^

We extend the contingency tables laid out previously (**Table 6**) to incorporate *β*, the relative risk of SARS-CoV-2 infection among vaccinated (vs. unvaccinated) individuals due to factors other than vaccination, as well as *ω*_*V*_ and *ω*_*U*_, the proportions of the vaccinated and unvaccinated populations, respectively, that remain uninfected. For simplicity, we assume near-total protection against reinfection conferred by naturally acquired immunity. Under the two designs,

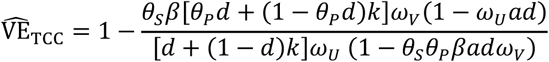

and

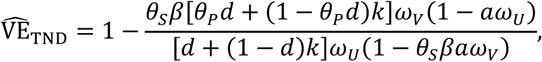

which, with low prevalence of infection (*a* ≈ 0) and low risk of disease due to other causes (*k* ≈ 0 or *k* ≪ *d*), yield

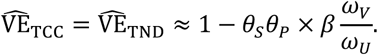

**Table 6:** Contingency table for TCC and TND studies with differential risk of current and prior infection among the vaccinated and unvaccinated.

| Exposure | Outcome |  |  |
| --- | --- | --- | --- |
|  | <u>Test-positive case (symptomatic)</u> | <u>Community control (asymptomatic)</u> | <u>Test-negative control (symptomatic)</u> |
| Vaccinated | $\nu \theta_s \beta a [\theta_p d + (1 - \theta_p d) k] \omega_v$ | $\nu (1 - k) (1 - \theta_s \theta_p \beta a d \omega_v)$ | $\nu (1 - \theta_s \beta a \omega_v) k$ |
| Unvaccinated | $(1 - \nu) a [d + (1 - d) k] \omega_u$ | $(1 - \nu) (1 - k) (1 - a d \omega_u)$ | $(1 - \nu) (1 - a \omega_u) k$ |

Here, differential risk of SARS-CoV-2 infection and prevalence of immunity among vaccinated and unvaccinated individuals lead to counteracting biases. With higher risk among individuals who receive vaccination (*β* > 1), we may expect higher prevalence of naturally acquired infection in this group and *ω*_*V*_ < *ω*_*U*_; bias is offset fully if *β*= *ω*_*U*_/*ω*_*V*_.

We plot conditions under which bias is driven predominantly by differential exposure to infection or by differential naturally acquired immunity in **Figure 4**. Considering the context of VE studies undertaken shortly after vaccine implementation, we may assume individuals’ time at risk is contributed primarily in the period before vaccination; with exponentially distributed infection times, and a cumulative force of infection *c* exerted over the period at risk, we may define the remaining uninfected populations *ω*_*U*_= exp (−*c*)and *ω*_*V*_= exp (−*βc*). Bias is fully offset with *c*= log(*β*)/(*β* − 1), yielding 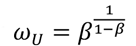 (**Figure 4**). Thus, the prevalence of pre-existing naturally acquired immunity needed to offset bias resulting from differential exposure is high, at 33% and 56% for *β* = 5 and *β* = 1.5, respectively.

**Figure 4:**
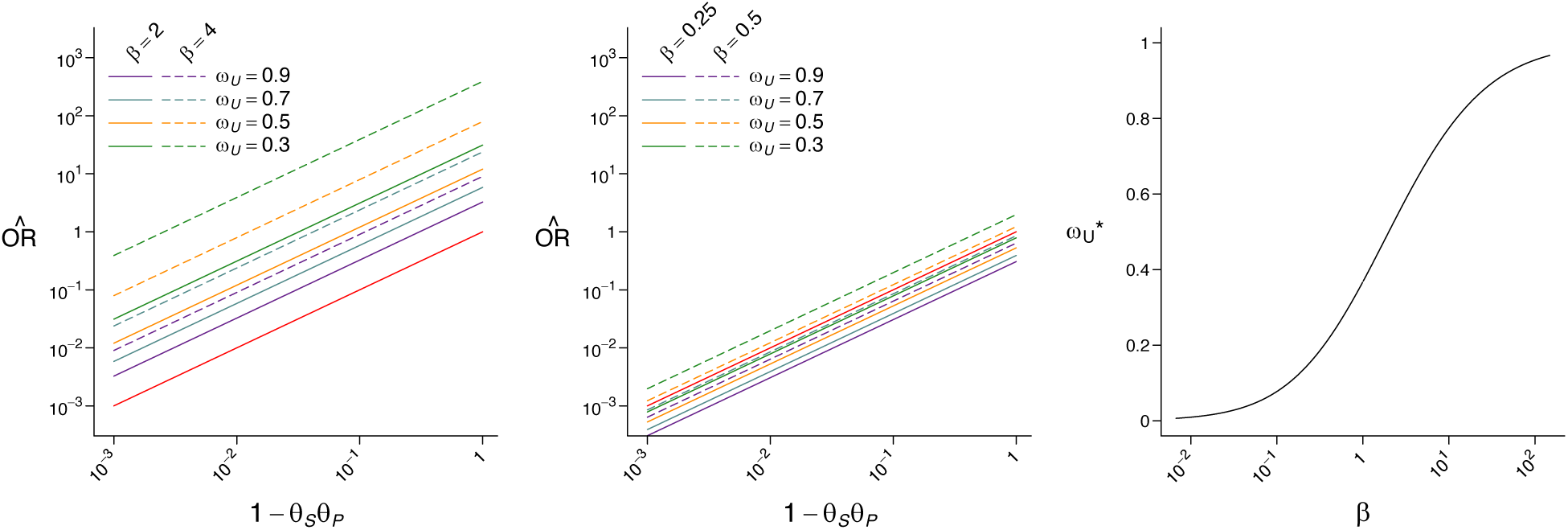
Estimated vaccine effectiveness under the TCC and TND with differential exposure to infection and prevalence of naturally acquired immunity among vaccinated and unvaccinated persons. We illustrate expected estimates of the odds ratio of vaccination given case versus control (or test-negative) status, under the limiting case of *a* ≈ 0 and *k* ≈ 0, under scenarios where vaccination occurs among individuals with higher (left panel: β *=* 2, solid lines; β *=* 4, dashed lines) or lower (center panel: β *=* 0.25, solid lines; β *=* 0.5, dashed lines) risk of exposure to SARS-CoV-2. Colors correspond to differing proportions of the unvaccinated population remaining susceptible to infection (*ω*_*U*_ *=* exp (−*c)*; we define the proportion of the vaccinated population remaining uninfected as *ω*_*V*_ *=* exp (−β*c)* for given values of *ω*_*U*_). Red diagonal lines across the left and center panels illustrate the true effect. The right panel illustrates threshold values of 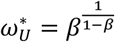at which the effects of differential exposure to SARS-CoV-2 and naturally-acquired immunity among vaccinated versus unvaccinated persons cancel out, resulting in a change in the direction of bias.

Detailed risk factor data are likely necessary to address confounding associated with differential risk of infection among individuals receiving and those not receiving vaccination. While certain occupations (e.g. healthcare worker or other essential worker) may be associated with heightened risk of exposure relative to the general public, substantial variation in risk may exist within these categories owing to differences in the specific tasks conducted and availability of as well as adherence to infection control measures. Individuals may modify aspects of their behavior following vaccine receipt, further altering differences in risk of infection among vaccine recipients and non-recipients. Collection of questionnaire items addressing risk behaviors and exposures is thus of importance to mitigate confounding. If it is possible to control for differences in individuals’ risk of infection, vaccine-unrelated differences in rates of infection and prevalence of natural immunity are not a prominent source of bias in the short term (*β* = 1 and *ω*_*U*_ = *ω*_*V*_).

### Temporal considerations

While we focus above on studies undertaken shortly after vaccine rollout, prior work has considered differential accrual of natural immunity among vaccinated and unvaccinated individuals over longer periods of time.^3,18,36,37^ For vaccines conferring “leaky” protection, TCC and TND studies may underestimate true protection owing to higher rates of acquisition of naturally-acquired immunity among unvaccinated individuals. This bias does not arise, however, for vaccines conferring “all-or-nothing” protection.^3,18^

## SUMMARY

Our analysis provides several practical insights to inform upcoming TCC and TND studies of SARS-CoV-2 vaccines. While we identify conditions and reasonable assumptions under which both designs can supply reliable VE estimates, several threats to validity should be taken into account in the design, analysis, and interpretation of TCC and TND studies. Below we offer considerations for researchers undertaking TCC and TND studies of vaccines against SARS-CoV-2:

### Use of severe COVID-19 clinical endpoints

Use of severe disease endpoints minimizes risk of misclassification and healthcare-seeking bias. To allow sufficient power, studies may need to enroll patients across multiple healthcare facilities, underscoring the need for standardized clinical criteria and case definitions. Studies addressing VE against common or less-severe clinical endpoints may benefit from using extended analytic methods that have been proposed previously for estimation of VE against diseases with non-specific diagnostic criteria.^11,38,39^ As these analysis methods must generally account for the strength of association between SARS-CoV-2 infection and clinical symptoms, enrollment and testing of control groups without symptoms related to SARS-CoV-2 infection may be of value for researchers studying VE against such endpoints.

### Use of molecular or antigen tests

Studies using SARS-CoV-2 detection assays with the highest analytic sensitivity may encounter the greatest risk of misclassifying symptomatic individuals at the end of their course of infection as cases, including when SARS-CoV-2 is not the cause of symptoms. This could be more problematic when prevalence of SARS-CoV-2 infection is high and outcomes of interest include non-severe or non-specific COVID-19 clinical manifestations, such as in outpatient settings. However, trade-offs associated with risk of bias must be balanced against assay specificity and test performance under varied prevalence and clinical settings. Where molecular assays with high analytical sensitivity are preferred for clinical care provision, researchers may consider analyzing quantitative cycle threshold data or collecting a second specimen for antigen-based testing.

### Collecting detailed information on clinical severity and testing indications

Reliance on clinic-based testing under both designs makes it difficult to identify and correct for differences in the likelihood for vaccinated and unvaccinated individuals, given infection and symptoms, to be ascertained as cases. Collection of standardized data on clinical symptoms among both cases and controls may reduce the risk of bias related to vaccine-associated differences in healthcare access or healthcare seeking. This strategy further provides an opportunity to identify and exclude, if applicable, registry-based or community controls who were ill but did not seek care. If testing is conducted for multiple pathogens, researchers may also investigate bias by assessing VE against other causes of illness not prevented by SARS-CoV-2 vaccination as a “sham” analysis to indicate risk of bias.^40,41,40,41^

### Background immunity

Prevalent naturally acquired immunity may complicate the interpretation of results under both the TCC and TND, potentially contributing to lower risk of infection and disease among individuals with the greatest risk of SARS-CoV-2 exposure. As initial vaccine doses will be prioritized to groups with high infection risk, individuals eligible to receive vaccine at the earliest opportunity may have higher prevalence of pre-existing naturally acquired immunity than those in lower-priority tiers. Studies may mitigate this bias, in part, by comparing individuals within the same prioritization groups, by collecting detailed risk factor data to control for variation in exposure, and through use of serological assays (if available) that differentiate vaccine-derived or naturally-induced responses (e.g. to non-vaccine antigens).

## Data Availability

This is a theoretical study without patient data; equations used to generate the figures appear in the manuscript.

---

**Table S1:** Contingency tables for TCC and TND studies with differing assay sensitivity for SARS-CoV-2 shedding and COVID-19.

| Exposure | Outcome |  |  |
| --- | --- | --- | --- |
|  | <u>Test-positive case</u><br>(symptomatic) | <u>Community control</u><br>(asymptomatic) | <u>Test-negative control (symptomatic)</u> |
| Vaccinated | $v\theta_s a[\theta_p d\sigma_p + (1 - \theta_p d)k\sigma_s]$ | $v(1 - k)(1 - \theta_s \theta_p ad)$ | $(1 - v)[k(1 - \theta_s a\sigma_s) + (1 - k)\theta_s \theta_p ad(1 - \sigma_p)]$ |
| Unvaccinated | $(1 - v)a[d\sigma_p + (1 - d)k\sigma_s]$ | $(1 - v)(1 - k)(1 - ad)$ | $(1 - v)[k(1 - a\sigma_s) + (1 - k)ad(1 - \sigma_p)]$ |

## Notes

**Sources of funding:** This work was supported by grants R01-AI14812701 from the National Institute for Allergy and Infectious Diseases to NPJ and JAL, and R01-AI139761 from the National Institute for Allergy and Infectious Diseases to NED.

### Competing Interest Statement

JAL has received grants and consulting fees from Pfizer, Inc. unrelated to this research.

### Funding Statement

This work was supported by grants R01-AI14812701 from the National Institute for Allergy and Infectious Diseases to NPJ and JAL, and R01-AI139761 from the National Institute for Allergy and Infectious Diseases to NED.

